# CURRENT TRENDS IN HOSPICE CARE USAGE FOR DIALYSIS PATIENTS IN THE USA

**DOI:** 10.1101/2023.05.30.23290755

**Authors:** Ayorinde I Soipe, John E Leggat, Ajibola I Abioye, Kriti Devkota, Fausat Oke, Kunal Bhuta, Moshood O Omotayo

## Abstract

**Background:** This study examined the predictors and latest trends in hospice utilization, adequate duration of hospice care, and dialysis discontinuation without hospice enrollment among patients with end stage renal disease (ESRD).

**Methods:** Data from the United States Renal Data System (USRDS) for ESRD patients who died between January 1, 2012, and December 31, 2019, were analyzed. Chi-square and logistic regression were used to evaluate associations between outcomes of interest and predictors while Joinpoint regression was used to examine trends.

**Results:** Among 803,049 patients, the median (IQR) age was 71 (17) years, 57% were male, 27% enrolled in hospice, 8% discontinued dialysis before death without hospice enrollment, and 7% remained in hospice for ≥15 days. Patients older than 64 years (adjusted odds ratio [aOR]: 2.75, 95% CI: 2.71-2.79) and white race (aOR: 1.79, 95% CI: 1.77-1.81) were more likely to enroll in hospice. White patients (aOR: 0.75, 95% CI: 0.73-0.76) and those who never received a kidney transplant (aOR: 0.75, 95% CI: 0.73-0.78) were less likely to have adequate duration of hospice care. Hospice enrollment and standardized duration of hospice care increased over time, with an average annual percentage change (AAPC) of 1.1% (95% CI:0.6 – 1.6) and 5% (95% CI:2.6 – 7.4) respectively.

**Conclusions:** Approximately one in every four ESRD patients who died between 2012 and 2019 had a history of hospice enrollment, while one in every 12 discontinued dialysis before death without hospice enrollment. There was an upward trend in the standardized duration of hospice care.

## INTRODUCTION

The increasing emphasis on value-based healthcare models nationally has created a renewed focus on hospice care as a holistic care model [1, 2]. Hospice care describes compassionate care at the end of life which involves expert medical care, pain management, emotional as well as spiritual support, all designed to meet patients’ specific wishes [3]. Hospice users among dialysis patients were less likely to die in the hospital, be hospitalized, admitted to the intensive care unit or undergo an intensive procedure in the last month of life and were associated with progressively lower healthcare costs compared to non-users, particularly for those initiating hospice care more than 15 days before death [4]. Despite this apparent benefit, while utilization of inpatient palliative care to address a spectrum of needs in ESRD and advanced kidney disease care has been on an upward trajectory, [5] the utilization of hospice care has been more complex. An analysis of hospice utilization between 2000 and 2014 showed that hospice utilization increased among ESRD patients; however, the rate of hospice use was lower when compared to patients with other life-limiting illnesses and length of stay was often less than 3 days [4].

The lower rates of hospice utilization and shorter length of stay among ESRD patients than among patients with other serious illnesses can be attributed to multiple barriers. A review noted that the need to stop dialysis before hospice admission, lack of knowledge among providers regarding hospice benefits for dialysis patients, and cultural considerations among patients are possible explanations for the low rate of hospice enrollment among ESRD patients [6]. Medicare policy, including payment models that forbid payment for dialysis by Medicare for patients with primary hospice diagnosis of ESRD, lack of hospice eligibility criteria specific to this population, and dialysis care quality metrics that are not aligned with quality of life constitute a structural barrier [7–10]. The either-or dichotomy of hospice and dialysis care that this policy fosters complicates decision-making regarding hospice utilization among ESRD patients, and it is widely regarded as the most important barrier. However, the fact that there is a sub-population that still does not access hospice care after discontinuing renal replacement therapy indicates that there are other significant barriers beyond the dilemma of having to choose between dialysis and hospice care induced by Medicare policy.

For hospice care to impact population-level ESRD or late-stage chronic kidney disease (CKD) outcomes, utilization patterns need to be improved. While few studies have examined sociodemographic and clinical correlates of hospice care among ESRD patients, those that have done so with a nationally representative population are rare, and there are no recent studies formally testing national trends in enrollment, duration of hospice care and dialysis discontinuation without hospice enrollment. Using data from the United States Renal Database System (USRDS), we sought to characterize trends and correlates of hospice enrollment in the ESRD population in the United States in the last decade. Specifically, we sought to answer three key questions: (i) what has been the trend and predictors of hospice enrollment? (ii) what has been the trend and predictors of hospice duration of care? (iii) what has been the trend in dialysis discontinuation without hospice enrollment?

## METHODS

### Data Source and Study Cohort

Using a retrospective observational study design, 3,473,037 unique patient records in the United States Renal Data System (USRDS) database were identified. The USRDS database is a national dataset containing information on diagnoses, demographic characteristics, clinical, biochemical, as well as claims information of ESRD patients [11]. Data reported in the Medical Evidence Form (CMS 2728) was utilized to assess outcome variables and covariates.

The final analytic sample comprised 803,049 unique records for ESRD patients who died between January 1, 2012, and December 31, 2019. The flowchart of records and how the final analytic sample was derived is shown in **Figure 1.** We chose 2012 as the cutoff because related analyses on initiation had been published in prior studies including a report sponsored by the Medicare Payment Advisory Commission, a United States government agency tasked with the role of advising the United States Congress on Medicare issues, which evaluated data up to 2012 [4, 12]. However, extended analyses for subsequent years were lacking despite changes in the national environment around value-based healthcare and policy initiatives being instituted. We chose 2019 cutoff because comprehensive data from USRDS was still being prepared from 2020 onwards at the time of our work. The institutional review board at Upstate Medical University approved the study protocol.

**Fig. 1.**
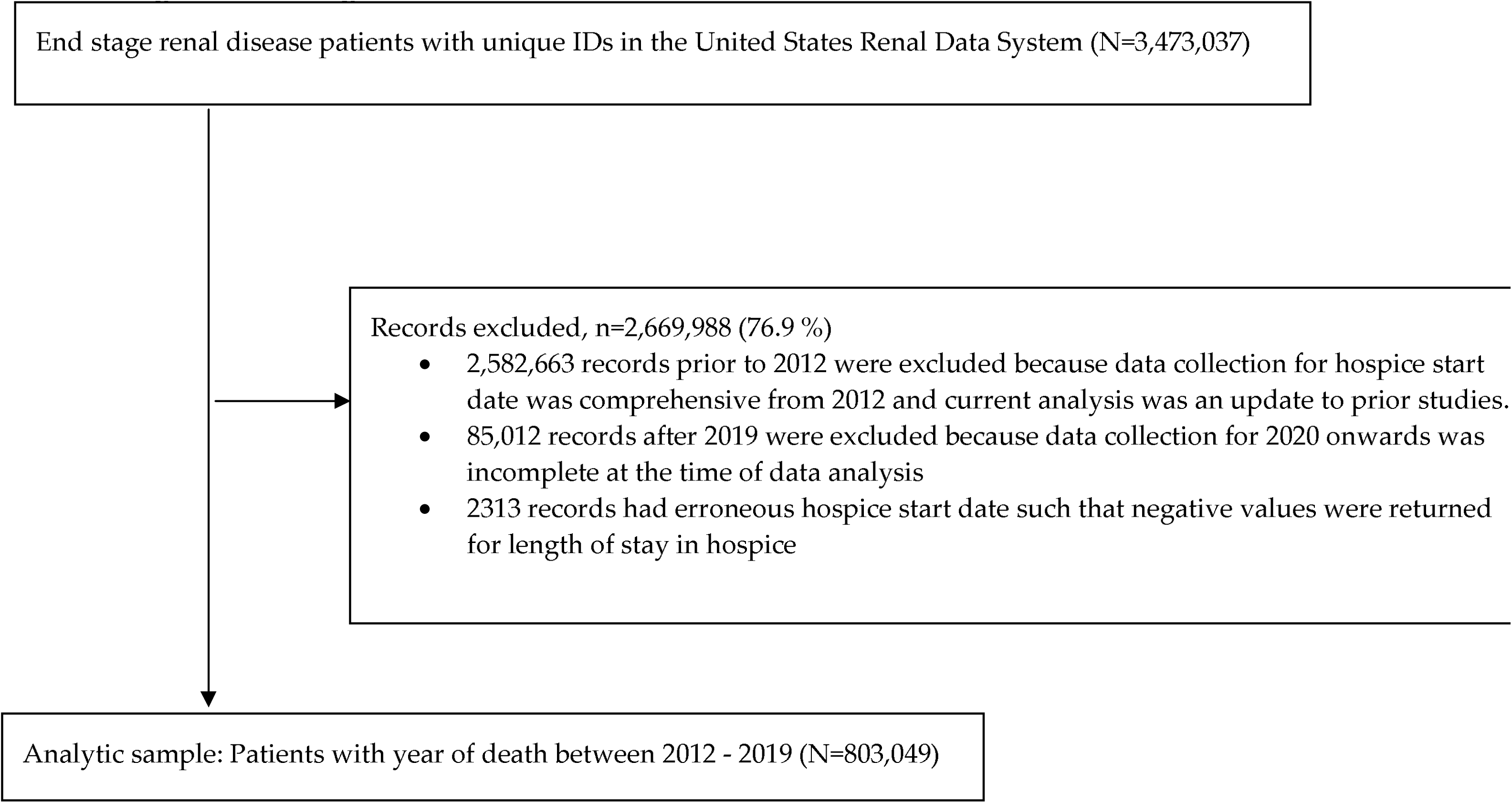
Flow diagram of cohort selection

### Outcomes

The outcomes of interest were hospice enrollment, duration of hospice care, and discontinuation of renal replacement therapy without hospice enrollment. The variable “hospice admit date” reported in the Medical Evidence Form (CMS 2728) was used to ascertain hospice enrollment, while the variable “renal replacement therapy discontinued prior to death” was used to define discontinuation of renal replacement therapy.

Duration of hospice care was dichotomized into adequate (15 days or more) and inadequate (less than 15 days), [4] while hospice enrollment was dichotomized into ‘yes vs no’ categories. Dialysis discontinuation without hospice enrollment was derived by combining the variables “hospice admit date” and “renal replacement therapy discontinued prior to death”, and thus reported as ‘yes’ for those who discontinued renal replacement therapy without hospice enrollment vs ‘not applicable’ for any other combination of responses.

### Covariates

Independent variables were identified *a priori* from the literature. These include age (≥ 65 years vs. < 65 years); gender (male vs. female); race (white vs. non-white); ethnicity (Hispanic vs. non-Hispanic); hypertension (yes vs. no); diabetes mellitus (yes vs. no); cancer (yes vs. no); COPD (yes vs. no); alcohol dependence (yes vs. no); drug dependence (yes vs. no); never received a kidney transplant (yes vs. no); non-renal congenital abnormality (yes vs. no). Newly created variables included macrovascular disease (yes vs. no) defined as history of atherosclerotic heart disease or peripheral vascular disease or congestive heart failure or myocardial infarction or ischemic heart disease or cerebrovascular disease or amputation; [13, 14] microvascular disease (yes vs. no) defined as history of diabetic retinopathy; [13, 14] debilitated (yes vs. no) defined as history of inability to transfer or inability to ambulate.

### Statistical Analysis

Descriptive statistics for categorical variables were reported as counts and proportions. Median and interquartile range [IQR] were calculated for continuous variables that were not normally distributed. In bivariate analysis, independent variables were categorized by hospice enrollment (yes vs no), dichotomized duration of hospice care (≥15 days vs <15 days), and Chi-square test was used to evaluate associations of the predictor variables with individual outcomes of interest. Estimates derived from the USRDS data were standardized to facilitate fair comparison of values derived from the cohort over time and to limit the effect of potential confounders which may be distributed differently in the population [15]. In multivariable analysis, logistic regression models were used to estimate adjusted odds ratios (ORs), with 95% confidence intervals (CI). The logistic regression model was evaluated using a backward selection procedure based on the Akaike Information Criterion (AIC). The model with the lowest AIC was considered the best fit. Joinpoint regression was utilized to compute the average annual percentage change (AAPC) in standardized duration of hospice care, standardized hospice enrollment proportion, as well as standardized proportion of dialysis discontinuation without hospice enrollment, in addition to being utilized to compare trends over time in standardized duration of hospice care between racial subgroups. The log-linear model processing option was selected while a permutation model selection method was utilized to estimate the AAPC in proportion and pairwise comparison of regression mean functions was used to compare the pace of change between racial subgroups. All primary statistical analysis was done using SAS software version 9.4 (SAS Institute, Cary, NC) and joinpoint regression was done using joinpoint software version 4.7.0.1 (Bethesda, MD).

### Missing data

For clinical comorbid conditions, 5.5% of the data were not recorded in the database for values that were not applicable to any particular patient. Data for key outcome variables utilized in statistical analysis were complete. Therefore, methods of missing data handling such as multiple imputation were not applied.

## RESULTS

There were 803,049 individuals in the analytic sample. The median age at the time of death was 71 years (interquartile range [IQR]: 62, 79); 64% were aged 65-89 years; 67.9% were white, and 57% were male. Among the analytic sample, 27% enrolled in hospice, 10% had a history of kidney transplantation, 7% had adequate duration of hospice care, and 8% discontinued dialysis prior to death without hospice enrollment (**Table 1**). Reasons attributed to dialysis discontinuation prior to death without hospice include failure of dialysis access, chronic failure to thrive, transplant failure, and following acute medical complication (**Online Resource 1**).

**Table 1:** Characteristics of United States Renal Data System (USRDS) patients with end stage renal disease (ESRD) who died between 2012 - 2019.

| Variable | n (%) |
| --- | --- |
| **Age at the time of death (N=803,049)** |  |
| <18 years | 707 (0.1) |
| 18-44 years | 32,150 (4.0) |
| 45-64 years | 222,448 (27.7) |
| 65-89 years | 515,726 (64.3) |
| >89 years | 32,018 (3.9) |
| **Age at the time of death in years (median, [IQR])** | 71 [62,79] |
| **Gender (N=803,049)** |  |
| Male | 456,172 (57) |
| Female | 346,977 (43) |
| **Race (N=803,049)** |  |
| White | 545,954 (67.9) |
| Black | 212,794 (26.5) |
| American Indian or Alaska Native | 8,081 (1.1) |
| Asian | 26,675 (3.3) |
| Native Hawaiian or Other Pacific Islander | 7,585 (0.9) |
| Other | 1,815 (0.2) |
| Unknown | 145 (0.02) |
| **Ethnicity (N=803,049)** |  |
| Hispanic | 102,952 (12.8) |
| Non-Hispanic | 695,707 (86.6) |
| Unknown | 4,390 (0.6) |
| **Never received a kidney transplant (N=803,049)** |  |
| Yes | 722,297 (90) |
| No | 80,752 (10) |
| **Enrolled in hospice (N=803,049)** |  |
| Yes | 218,635 (27) |
| No | 584,414 (73) |
| **Duration of hospice care in days (N=803,049)** |  |
| ≥15 days | 54,990 (7) |
| <15 days | 163,645 (20) |
| Not applicable / non hospice users | 584,414 (73) |
| Median, [IQR] | 5, [2,15] |
| **Dialysis discontinued prior to death without hospice enrollment (N=803,049)** |  |
| Yes | 66,209 (8) |
| Not applicable | 736,840 (92) |

In unadjusted analysis: age, gender, race, ethnicity, hypertension, diabetes mellitus, debilitation, cancer, COPD, macrovascular disease, microvascular disease, alcohol dependence, drug dependence, never receiving a kidney transplant and non-renal congenital abnormality were associated with hospice enrollment. Similarly, gender, race, ethnicity, diabetes mellitus, debilitation, macrovascular disease, alcohol dependence, drug dependence, and never receiving a kidney transplant were associated with duration of hospice care ≥15 days (**Table 2)**.

**Table 2:** Characteristics of United States Renal Data System (USRDS) patients with end stage renal disease (ESRD) who died between 2012 and 2019 categorized by hospice enrollment status and duration of hospice care

| Variable | Enrolled in hospice |  |  |  | Duration of hospice care |  |  |  |
| --- | --- | --- | --- | --- | --- | --- | --- | --- |
|  | Overall count (n, %) | Yes | No | p value | Overall count (n, %) | ≥15 days | <15 days | p value |
| **Age (years)** | (N=803,049) |  |  | <0.001 | (N=218,635) |  |  | 0.70 |
| ≥ 65 (n, %) | 547,744 (68) | 182,428 (33) | 365,316 (67) |  | 182,428 (84) | 45,912 (25) | 136,516 (75) |  |
| < 65 (n, %) | 255,305 (32) | 36,207 (14) | 219,098 (86) |  | 36,207 (16) | 9,078 (25) | 27,129 (75) |  |
| **Gender** | (N=803,049) |  |  | <0.001 | (N=218,635) |  |  | <0.001 |
| Male (n, %) | 456,172 (57) | 119,519 (26) | 336,653 (74) |  | 119,519 (55) | 29,686 (25) | 89,833 (75) |  |
| Female (n, %) | 346,877 (43) | 99,116 (29) | 247,761 (71) |  | 99,116 (45) | 25,304 (25) | 73,812 (75) |  |
| **Race** | (N=803,049) |  |  | <0.001 | (N=218,635) |  |  | <0.001 |
| White (n, %) | 545,954 (68) | 168,353 (31) | 377,601 (69) |  | 168,353 (77) | 40,464 (24) | 127,892 (76) |  |
| Non-white (n, %) | 257,095 (32) | 50,282 (20) | 206,813 (80) |  | 50,282 (23) | 14,529 (29) | 35,753 (71) |  |
| **Ethnicity** | (N=798,659) |  |  | <0.001 | (N=217,309) |  |  | <0.001 |
| Hispanic (n, %) | 102,952 (13) | 22,157 (22) | 80,795 (78) |  | 22,157 (10) | 6,274 (28) | 15,883 (72) |  |
| Non-Hispanic (n, %) | 695,707 (87) | 195,152 (28) | 500,555 (72) |  | 195,152 (90) | 48,228 (25) | 146,924 (75) |  |
| **Clinical comorbid conditions** |  |  |  |  |  |  |  |  |
| **Hypertension** | (N=758,717) |  |  | <0.001 | (N=207,438) |  |  | 0.09 |
| Yes (n, %) | 667,186 (88) | 181,854 (27) | 485,332 (73) |  | 181,854 (88) | 45,461 (25) | 136,393 (75) |  |
| No (n, %) | 91,531 (12) | 25,584 (28) | 65,947 (72) |  | 25,584 (12) | 6,243 (24) | 19,311 (76) |  |
| **Diabetes mellitus** | (N=758,717) |  |  | <0.001 | (N=207,438) |  |  | <0.001 |
| Yes (n, %) | 433,091 (57) | 110,437 (26) | 322,654 (74) |  | 110,437 (53) | 26,964 (24) | 83,473 (76) |  |
| No (n, %) | 325,626 (43) | 97,001 (30) | 228,625 (70) |  | 97,001 (47) | 24,770 (26) | 72,231 (74) |  |
| **Debilitated** | (N=758,717) |  |  | <0.001 | (N=207,438) |  |  | <0.001 |
| Yes (n, %) | 63,575 (8) | 18,377 (29) | 45,198 (71) |  | 18,377 (9) | 4,986 (27) | 13,397 (73) |  |
| No (n, %) | 695,142 (92) | 189,061 (27) | 506,081 (73) |  | 189,061 (91) | 46,768 (25) | 142,313 (75) |  |
| **Cancer** | (N=758,717) |  |  | <0.001 | (N=207,438) |  |  | 0.39 |
| Yes (n, %) | 59,478 (8) | 22,951 (39) | 36,527 (61) |  | 22,951 (11) | 5,776 (25) | 17,175 (75) |  |
| No (n, %) | 699,239 (92) | 184,487 (26) | 514,752 (74) |  | 184,487 (89) | 45,958 (25) | 138,529 (75) |  |
| **COPD** | (N=758,717) |  |  | <0.001 | (N=207,438) |  |  | 0.61 |
| Yes (n, %) | 80,412 (11) | 24,403 (30) | 56,009 (70) |  | 24,403 (12) | 6,054 (25) | 18,349 (75) |  |
| No (n, %) | 678,305 (89) | 183,035 (27) | 495,270 (73) |  | 183,035 (88) | 45,680 (25) | 137,355 (75) |  |
| **Macrovascular disease** | (N= 758,717) |  |  | <0.001 | (N=207,438) |  |  | <0.001 |
| Yes (n, %) | 384,410 (51) | 106,993 (28) | 277,417 (72) |  | 106,993 (52) | 26,334 (25) | 80,659 (75) |  |
| No (n, %) | 374,307 (49) | 100,445 (27) | 273,862 (73) |  | 100,445 (48) | 25,400 (25) | 75,045 (75) |  |
| **Microvascular disease** | (N=758,717) |  |  | <0.001 | (N=207,438) |  |  | 0.41 |
| Yes (n, %) | 55,120 (7) | 11,783 (21) | 43,337 (79) |  | 11,783 (6) | 2,976 (25) | 8,807 (75) |  |
| No (n, %) | 703,597 (93) | 195,655 (28) | 507,942 (72) |  | 195,655 (94) | 48,758 (25) | 146,897 (75) |  |
| **Alcohol dependence** | (N=758,717) |  |  | <0.001 | (N=207,438) |  |  | 0.001 |
| Yes (n, %) | 12,604 (2) | 2,773 (22) | 9,831 (78) |  | 2,773 (1) | 764 (28) | 2,009 (72) |  |
| No (n, %) | 746,113 (98) | 204,665 (27) | 541,448 (73) |  | 204,665 (99) | 50,970 (25) | 153,695 (75) |  |
| Drug dependence | (N=758,717) |  |  | <0.001 | (N=207,438) |  |  | <0.001 |
| Yes (n, %) | 9,731 (1) | 1,286 (13) | 8,445 (87) |  | 1,286 (0.6) | 381 (30) | 905 (70) |  |
| No (n, %) | 748,986 (99) | 206,152 (27) | 542,834 (73) |  | 206,152 (99.4) | 51,353 (25) | 154,799 (75) |  |
| Never received a kidney transplant | (N=803,049) |  |  | <0.001 | (N=218,635) |  |  | <0.001 |
| Yes (n, %) | 722,297 (90) | 199,021 (28) | 523,276 (72) |  | 199,021 (91) | 49,107 (25) | 149,914 (75) |  |
| No (n, %) | 80,752 (10) | 19,614 (24) | 61,138 (76) |  | 19,614 (9) | 5,883 (30) | 13,731 (70) |  |
| Non-renal congenital abnormality | (N=758,717) |  |  | <0.001 | (N=207,438) |  |  | 0.79 |
| Yes (n, %) | 1,340 (0.2) | 272 (20) | 1,068 (80) |  | 272 (0.1) | 66 (24) | 206 (76) |  |
| No (n, %) | 757,377 (99.8) | 207,166 (27) | 550,211 (73) |  | 207,166 (99.9) | 51,668 (25) | 155,498 (75) |  |

p-values were obtained from chi square test

In logistic regression models controlling for clinical and sociodemographic characteristics, the model with the lowest AIC was selected as the best fit. In the final models, patients who are older than 64 years (adjusted odds ratio [aOR]: 2.75, 95% CI: 2.71-2.79), those who never received a kidney transplant (aOR: 1.07, 95% CI: 1.05-1.09), and those who are of the white race (aOR: 1.79, 95% CI: 1.77-1.81) were more likely to enroll in hospice, while patients who are white (aOR: 0.75, 95% CI: 0.73-0.76), and those who never received a kidney transplant (aOR: 0.75, 95% CI: 0.73-0.78) were less likely to have adequate duration of hospice care (**Table 3)**.

**Table 3:** Logistic regression modeling of hospice enrollment and adequate duration of hospice care of stay among United States Renal Data System (USRDS) patients with end stage renal disease (ESRD) who died between 2012 and 2019.

| **Characteristic** | **<u>Enrolled in hospice</u>**<br>**aOR (95% C.I)** | **<u>Adequate duration of hospice care</u>**<br>**aOR (95% C.I)** |
| --- | --- | --- |
| **Age (years)** |  |  |
| ≥ 65 (ref: < 65) | 2.75 (2.71 – 2.79) | - |
| **Gender** |  |  |
| Female (ref: Male) | 1.16 (1.15 – 1.17) | 1.03 (1.01 – 1.05) |
| **Race** |  |  |
| White (ref: Non-white) | 1.79 (1.77 – 1.81) | 0.75 (0.73 – 0.76) |
| **Ethnicity** |  |  |
| Non-Hispanic (ref: Hispanic) | 1.55 (1.52 – 1.57) | 0.77 (0.74 – 0.79) |
| **Clinical comorbid conditions** |  |  |
| Cancer: yes (ref: no) | 1.42 (1.39 – 1.45) | - |
| Alcohol dependence: yes (ref: no) | 1.04 (0.99 – 1.08) | 1.09 (1.01 – 1.19) |
| Never received a kidney transplant: yes (ref: no) | 1.07 (1.05 – 1.09) | 0.75 (0.73 – 0.78) |
| Debilitated: yes (ref: no) | 1.03 (1.01 – 1.05) | 1.16 (1.12 – 1.19) |
| Hypertension: yes (ref: no) | 1.01 (0.99 – 1.03) | - |
| COPD: yes (ref: no) | 0.98 (0.97 – 0.99) | - |
| Macrovascular disease: yes (ref: no) | 0.94 (0.93 – 0.95) | - |
| Microvascular disease: yes (ref: no) | 0.88 (0.86 – 0.90) | - |
| Non-renal congenital abnormality: yes (ref: no) | 0.87 (0.76 – 0.99) | - |
| Diabetes mellitus: yes (ref: no) | 0.86 (0.85 – 0.87) | 0.93 (0.91 – 0.95) |
| Drug dependence: yes (ref: no) | 0.75 (0.70 – 0.79) | 1.15 (1.02 – 1.30) |

aOR = adjusted odds ratio; CI=confidence interval
Adjusted odds ratios were obtained from logistic regression via a backward selection modeling procedure

Standardized proportion of patients who discontinue renal replacement therapy every year between 2012 and 2019 without hospice enrollment remained relatively constant at approximately 11% per year with a non-significant trend towards a decrease over time (AAPC - 0.4%, 95% CI: −1.2 to 0.3) (**Table 4)**. However, standardized hospice enrollment proportion increased over time (AAPC 1.1%, 95% CI: 0.6 to 1.6) and standardized duration of hospice care increased over time (AAPC 5%, 95% CI: 2.6 to 7.4). Comparing trends among racial subgroups, change in standardized duration of hospice care among black patients was significantly higher (AAPC: 7.6, 95% CI: 3.9-11.4) compared to White (AAPC: 4.0, 95% CI: 1.9-6.1) and Asian (AAPC: 6.6, 95% CI: 2.0-11.4) patients (**Table 4**).

**Table 4:** Trends in standardized duration of hospice care, hospice enrollment, and dialysis discontinuation without hospice enrollment, among end stage renal disease (ESRD) patients categorized by year of death [2012–2019]

|  | 2012 | 2013 | 2014 | 2015 | 2016 | 2017 | 2018 | 2019 | AAPC:<br>% (95% CI) | p value |
| --- | --- | --- | --- | --- | --- | --- | --- | --- | --- | --- |
| <sup>a</sup> Standardized duration of hospice care (days) | 22.7 | 25.9 | 28.7 | 30.2 | 32.4 | 32.5 | 33.7 | 32.4 | 5.0* (2.6 to 7.4) | <0.001 |
| <sup>b</sup> Standardized proportion of hospice enrollment (%) | 26.3 | 26.8 | 26.9 | 26.9 | 27.2 | 27.1 | 27.8 | 28.6 | 1.1* (0.6 to 1.6) | <0.001 |
| <sup>c</sup> Standardized proportion of dialysis discontinuation without hospice enrollment (%) | 11.7 | 11.3 | 11.1 | 11.7 | 11.3 | 11.1 | 11.1 | 11.3 | -0.4 (-1.2 to 0.3) | 0.22 |
| <sup>d</sup> Standardized duration of hospice care by race (days) |  |  |  |  |  |  |  |  |  |  |
| American Indian | 15.1 | 22.2 | 26.2 | 23.8 | 39.9 | 48.5 | 22.0 | 23.8 | 3.3 (-11.1 to 19.9) | 0.67 |
| Asian | 22.1 | 22.8 | 30.5 | 33.4 | 28.5 | 36.7 | 32.5 | 34.8 | 6.6* (2.0 to 11.4) | 0.01 |
| Black | 27.8 | 34.3 | 36.7 | 40.7 | 46.7 | 52.1 | 54.8 | 45.9 | 7.6* (3.9 to 11.4) | <0.001 |
| Other | 25.3 | 17.7 | 14.3 | 35.1 | 72.1 | 13.8 | 92.2 | 60.5 | 19.5 (-6.9 to 53.4) | 0.13 |
| Native Hawaiian | 23.7 | 28.9 | 16.4 | 25.9 | 32.6 | 21.1 | 28.8 | 42.6 | 6.2 (-3.9 to 17.5) | 0.19 |
| White | 21.5 | 23.9 | 25.9 | 27.5 | 28.8 | 27.3 | 28.5 | 28.8 | 4.0* (1.9 to 6.1) | <0.001 |

<sup>a</sup> Standardized duration of hospice care = summation of duration of hospice care contributed by subjects per year / number of hospice enrollment and death for each year
<sup>b</sup> Standardized % = [number of hospice enrollment and death for each year/ number of deaths per year] \* 100
<sup>c</sup> Standardized % = [number who died but discontinued dialysis prior to death without hospice enrollment for each year/ number who died without hospice enrollment per year] \* 100
<sup>d</sup> Standardized duration of hospice care by race = summation of duration of hospice care contributed by subjects in each race and year per cell / number of hospice enrollment and death for each race and year per cell
AAPC = average annual percentage change
\* Implies significant finding.
Estimates were reported consistently as single decimal place for log-linear processing in joinpoint regression
Proportion of sample cohorts utilized for standardization are presented in the Online Resource 2

## DISCUSSION

This study examined the sociodemographic and clinical correlates of hospice enrollment and duration of hospice care as well as trends in dialysis discontinuation among ESRD patients in the United States who died between 2012 and 2019. We found that though hospice enrollment was common, a meaningful proportion of patients discontinued renal replacement therapy without hospice enrollment. The median duration of hospice care was 5 days and standardized duration of hospice care increased over time. Elderly patients, White patients, and patients who never received a kidney transplant were more likely to enroll in hospice.

Hospice utilization and duration of hospice care among the ESRD population in the United States is an area of investigation that has been garnering increasing attention given the mounting evidence of the value of hospice care [16, 17]. Hospice utilization among ESRD decedents was estimated to be 20% in the study published by Watcherman et al based on USRDS data [4] This estimate shows a lower rate of hospice utilization among ESRD patients when compared to patients with other chronic medical conditions. The lower rate of hospice utilization among ESRD patients can be attributed to certain unique barriers such as the requirement to stop dialysis before hospice admission [6, 16]. In our analysis, we showed that hospice utilization had increased since the analysis done by Watcherman et al [4]. This is also in keeping with the findings from prior research which had shown a steady but slight increase in hospice utilization among ESRD patients as far back as year 2000 [18]. Furthermore, we also showed that those who did not enroll in hospice after dialysis discontinuation has however been constant. With this trend, interventions for improving care coordination and knowledge and communication skills about hospice care among kidney care providers are needed.

In our analysis, the median duration of hospice care was 5 days with an increase in standardized duration of hospice care over the study period. This is similar to the hospice length of stay among ESRD patients within the USRDS database who died between 2000 - 2014 published in the analysis by Wachterman et al., [4] while we further showed that there has been a significant increase in standardized duration of hospice care from 2012 – 2019. Furthermore, approximately 1 in every dozen ESRD patients discontinued renal replacement therapy prior to death and did not enroll in hospice. It is important to note that the terms “dialysis discontinuation before death” and “withdrawal from dialysis” are historically not considered to be the same. While “withdrawal from dialysis” was once coded as a cause of death in the death notification forms of patients with ESRD, “dialysis discontinuation” was not considered to be a surrogate term by many nephrologists [19–21]. Furthermore, debate remains ongoing on how the terms should be appropriately defined [22]. Historically, studies had shown that between 6 – 22% of ESRD patients withdraw from dialysis [23–25]. More recently, Wetmore et al estimated that approximately 9% of patients who died between 2010 and 2015 electively withdrew from dialysis [20]. Various factors that have been associated with dialysis withdrawal include older age, female gender, white race, being from a rural setting, as well as factors important to physical independence including history of cerebrovascular disease [20, 26].

In our analysis, we showed that 8% of all ESRD patients who died between 2012 and 2019 had discontinued renal replacement therapy prior to death without hospice enrollment. More importantly, our analysis highlighted that the standardized proportion of ESRD patients who discontinue renal replacement therapy every year between 2012 and 2019 without hospice enrollment remained relatively constant at approximately 11% per year. In our sample, the reasons reported as being associated with dialysis discontinuation prior to death among those who did not enroll in hospice include failure of dialysis access, chronic failure to thrive, transplant failure, and following acute medical complication. Patients who discontinue dialysis without hospice care are an important subset of ESRD patients that may benefit from better care coordination between organizations and professionals providing advanced kidney care and hospice care, thereby improving hospice enrollment and adequate duration of hospice care. These metrics have been demonstrated in prior studies to be associated with better value of care [27]. Therefore, policies that would encourage continuation of dialysis for comfort and symptom management at the end of life should be promoted.

The newly introduced Medicare Kidney Care Choices (KCC) model, if expanded, promises to be a remedy for the structural constraints that current Medicare Hospice policy poses to hospice utilization in late-stage CKD and ESRD care [28]. The KCC, particularly its Comprehensive Kidney Care Contracting (CKCC) global option, can incentivize nephrology practices and dialysis centers to take appropriate advantage of the holistic care and value that hospice care can provide. This model is based on the global population-based payment option for improving hospice utilization rates and duration of hospice care in this population.

A strength of our study is the use of a national dataset with adequate sample sizes that allowed formal testing of national trends, and a combination of important socio-demographic and clinical predictors in our models. Also, our analysis is timely considering the commencement of the Medicare KCC model demonstration in 2022. However, our conclusions should be interpreted with caution. One limitation of our study is missing data for comorbidities, but it is likely that the mechanism of missingness is random. Yet, the potential influence of the missingness should be considered when interpreting the results.

Secondly, the results should further be interpreted with caution due to the retrospective design of the study whereby data captured in the database and analyzed can only be assumed to be accurate. Thirdly, our analysis did not include years beyond 2019. The literature is awash with studies which examined the impact of the COVID-19 pandemic on healthcare services across the globe and across disciplines [29, 30]. However, the trajectory of how the pandemic has affected hospice utilization patterns in this specific population remains lacking.

In summary, approximately 1 in every 4 ESRD patient who died between 2012 – 2019 had a history of hospice enrollment while 1 in every 12 discontinued renal replacement therapy prior to death and did not enroll in hospice. Among ESRD decedents with hospice enrollment, the median duration of hospice care before death was 5 days. Elderly patients, White patients, and patients who never received a kidney transplant were more likely to enroll in hospice, and standardized duration of hospice care saw an upward trend. It is crucial to enhance coordination between organizations and professionals involved in ESRD and hospice care. Patients who discontinue dialysis without hospice enrollment will likely require different interventions to improve hospice enrollment beyond policy interventions to promote concurrent hospice/dialysis care. Once a patient decides to stop dialysis, the Medicare barrier to hospice enrollment ought to readily dissolve, so there must be other factors impeding these patients from enrolling in hospice. This observation necessitates further exploration.

[FIG]

## ONLINE RESOURCE FIGURE AND TABLE LEGEND

**Online Resource 1**: Reasons attributed to dialysis discontinuation prior to death among those who did not enroll in hospice within the sample of end-stage renal disease (ESRD) patients who died between 2012 – 2019.

**Online Resource 2**: Proportion of sample cohorts by year of death

## Statements and Declarations

### Data sharing statement and Code Availability

The data analyzed for this study was from a publicly available database. The deidentified participant data, including demographic information, the study outcomes (hospice enrollment, duration of hospice care, and dialysis discontinuation without hospice enrollment), and other covariables, will be shared after the study results are published. To request access to this information, individuals should contact Dr. Ayorinde Soipe. Data access must be approved by the Upstate Medical University Institutional Review Board and National Institute of Diabetes and Digestive and Kidney Diseases Coordinating Center. The SAS codes utilized for analysis and estimates derived from the analysis will be provided Dr. Ayorinde Soipe upon request.

### Disclaimer

The data reported here have been supplied by the United States Renal Data System (USRDS). The interpretation and reporting of these data are the responsibility of the author(s) and in no way should be seen as an official policy or interpretation of the United States government.

### Author contributions

Conceptualization: Ayorinde Soipe, Omotayo Moshood; Methodology: Ayorinde Soipe, Omotayo Moshood, Ajibola Abioye; Formal analysis and interpretation: Ayorinde Soipe, Ajibola Abioye, Omotayo Moshood, John Leggat; Writing - original draft preparation: Ayorinde Soip, Omotayo Moshoode; Writing - review and editing: Ayorinde Soipe, John Leggat, Ajibola Abioye, Kriti Devkota, Fausat Oke, Kunal Bhuta, Moshood Omotayo; Resources: Kriti Devkota; Supervision: John Leggat

## Supporting information

Online resources

## Acknowledgments

We acknowledge Dr. Debra Luczkiewicz, MD for her expertise and assistance.

## REFERENCES

[1] G. Jain and D. E. Weiner, “Value-based care in nephrology: The Kidney Care Choices Model and other reforms,” Kidney360, vol. 2, no. 10, p. 1677, 2021.

[2] S. L. Tummalapalli and M. L. Mendu, “Value-Based Care and Kidney Disease: Emergence and Future Opportunities,” Advances in Chronic Kidney Disease, vol. 29, no. 1, pp. 30–39, 2022.

[3] D. Casey, “Hospice and Palliative Care: What’s the Difference?,” Medsurg Nursing, vol. 28, no. 3, pp. 196–197, 2019.

[4] M. W. Wachterman, S. M. Hailpern, N. L. Keating, M. K. Tamura, and A. M. O’Hare, “Association between hospice length of stay, health care utilization, and Medicare costs at the end of life among patients who received maintenance hemodialysis,” JAMA internal medicine, vol. 178, no. 6, pp. 792–799, 2018.

[5] Y. Wen, et al., “Trends and racial disparities of palliative care use among hospitalized patients with ESKD on dialysis,” Journal of the American Society of Nephrology, vol. 30, no. 9, pp. 1687–1696, 2019.

[6] N. R. O’Connor, “Hospice Among Hemodialysis Patients: Too Little, Too Late to Impact Care Delivery or Costs?,” American Journal of Kidney Diseases, vol. 72, no. 6, pp. 903–905, 2018.

[7] M. K. Tamura and D. E. Meier, “Five policies to promote palliative care for patients with ESRD,” Clinical Journal of the American Society of Nephrology, vol. 8, no. 10, pp. 1783–1790, 2013.

[8] J. O. Schell and D. S. Johnson, “Challenges with providing hospice care for patients undergoing long-term dialysis,” Clinical Journal of the American Society of Nephrology, vol. 16, no. 3, pp. 473–475, 2021.

[9] D. D. Trivedi, “Palliative dialysis in end-stage renal disease,” American Journal of Hospice and Palliative Medicine®, vol. 28, no. 8, pp. 539–542, 2011.

[10] D. Castner and D. Bednarski, “The intersection of the Medicare end-stage renal disease (ESRD) benefit and hospice: an overview for home care and hospice clinicians,” Home Healthcare Now, vol. 29, no. 8, pp. 464–476, 2011.

[11] United States Renal Data System (USRDS). 2020 USRDS annual data report: Epidemiology of kidney disease in the United States. National Institutes of Health, National Institute of Diabetes and Digestive and Kidney Diseases, Bethesda, MD, 2020. [Online] Available: https://www.niddk.nih.gov/about-niddk/strategic-plans-reports/usrds

[12] C. Hogan and M. P. A. Commission, “Spending in the last year of life and the impact of hospice on Medicare outlays,” A report by Direct Research, LLC, for the Medicare Payment Advisory Commission, 2015.

[13] R. E. Climie, et al., “Macrovasculature and microvasculature at the crossroads between type 2 diabetes mellitus and hypertension,” Hypertension, vol. 73, no. 6, pp. 1138–1149, 2019.

[14] K. C. Donaghue, F. Chiarelli, D. Trotta, J. Allgrove, and K. Dahl-Jorgensen, “Microvascular and macrovascular complications,” ed: Blackwell Publishing Ltd Oxford, UK, 2007.

[15] R. Farmer, R. Lawrenson, and D. Miller, Epidemiology and public health medicine. Blackwell, 2004.

[16] K. F. Thompson, J. Bhargava, R. Bachelder, R. Bova-Collis, and A. H. Moss, “Hospice and ESRD: knowledge deficits and underutilization of program benefits,” Nephrol Nurs J, vol. 35, no. 5, pp. 461–505, 2008.

[17] M. W. Wachterman, E. E. Corneau, A. M. O’Hare, N. L. Keating, and V. Mor, “Association of Hospice Payer With Concurrent Receipt of Hospice and Dialysis Among US Veterans With End-stage Kidney Disease: A Retrospective Analysis of a National Cohort,” in JAMA Health Forum, 2022, vol. 3, no. 10: American Medical Association, pp. e223708–e223708.

[18] A. M. Murray, C. Arko, S.-C. Chen, D. T. Gilbertson, and A. H. Moss, “Use of hospice in the United States dialysis population,” Clinical Journal of the American Society of Nephrology, vol. 1, no. 6, pp. 1248–1255, 2006.

[19] J. L. Holley, “A single-center review of the death notification form: discontinuing dialysis before death is not a surrogate for withdrawal from dialysis,” American journal of kidney diseases, vol. 40, no. 3, pp. 525–530, 2002.

[20] J. B. Wetmore, H. Yan, D. T. Gilbertson, and J. Liu, “Factors Associated with Elective Withdrawal of Maintenance Hemodialysis: A Case-Control Analysis,” American journal of nephrology, vol. 51, no. 3, pp. 227–236, 2020.

[21] E. Murphy, M. J. Germain, H. Cairns, I. J. Higginson, and F. E. Murtagh, “International variation in classification of dialysis withdrawal: a systematic review,” Nephrology Dialysis Transplantation, vol. 29, no. 3, pp. 625–635, 2014.

[22] J. H. Chen, W. H. Lim, and P. Howson, “Changing landscape of dialysis withdrawal in patients with kidney failure: Implications for clinical practice,” Nephrology, 2022.

[23] J. Leggat, W. E. Bloembergen, G. Levine, T. E. Hulbert-Shearon, and F. K. Port, “An analysis of risk factors for withdrawal from dialysis before death,” Journal of the American Society of Nephrology, vol. 8, no. 11, pp. 1755–1763, 1997.

[24] J. E. Leggat Jr, R. D. Swartz, and F. K. Port, “Withdrawal from dialysis: a review with an emphasis on the black experience,” Advances in renal replacement therapy, vol. 4, no. 1, pp. 22–29, 1997.

[25] H. A. Qazi, H. Chen, and M. Zhu, “Factors influencing dialysis withdrawal: a scoping review,” BMC nephrology, vol. 19, no. 1, pp. 1–11, 2018.

[26] J. A. Hussain, K. Flemming, F. E. Murtagh, and M. J. Johnson, “Patient and health care professional decision-making to commence and withdraw from renal dialysis: a systematic review of qualitative research,” Clinical Journal of the American Society of Nephrology, vol. 10, no. 7, pp. 1201–1215, 2015.

[27] K. Romero, E. Widera, and M. W. Wachterman, “Breaking the Link Between Enrollment in Hospice and Discontinuation of Dialysis,” JAMA Internal Medicine, 2023.

[28] A. V. Kshirsagar et al., “Keys to Driving Implementation of the New Kidney Care Models,” Clinical Journal of the American Society of Nephrology, 2022.

[29] T. K. Novick, K. Rizzolo, and L. Cervantes, “COVID-19 and Kidney Disease Disparities in the United States,” (in eng), Adv Chronic Kidney Dis, vol. 27, no. 5, pp. 427–433, Sep 2020, doi: 10.1053/j.ackd.2020.06.0051.

[30] R. Moynihan, et al., “Impact of COVID-19 pandemic on utilisation of healthcare services: a systematic review,” BMJ open, vol. 11, no. 3, p. e045343, 2021.

