## Supplementary material for "CURRENT TRENDS IN HOSPICE CARE USAGE FOR DIALYSIS PATIENTS IN THE USA": Online resources

ONLINE RESOURCE FIGURE AND TABLE LEGEND

Online Resource 1: Reasons attributed to dialysis discontinuation prior to death among those who did not enroll in hospice within the sample of end-stage renal disease (ESRD) patients who died between 2012 – 2019.

Online Resource 2: Proportion of sample cohorts by year of death

**Online Resource 1: Reasons attributed to dialysis discontinuation prior to death among those who did not enroll in hospice within the sample of end-stage renal disease (ESRD) patients who died between 2012 – 2019 (N=66,209).**

| **Reason** | **Count (n, %)** |
| --- | --- |
| Following hemodialysis and/or peritoneal dialysis access failure | 31 (0.05) |
| Following transplant failure | 6 (0.01) |
| Following chronic failure to thrive | 1,028 (1.55) |
| Following acute medical complication | 886 (1.34) |
| Other | 691 (1.04) |
| Dialysis stop reason unknown | 63,567 (96.01) |

**Online Resource 2: Proportion of sample cohorts by year of death**

| **Variable** | n (%) |
| --- | --- |
| **Year of death for the entire cohort with or without hospice enrollment (N=803,049)** |  |
| 2012 | 90,895 (11) |
| 2013 | 92,466 (12) |
| 2014 | 95,177 (12) |
| 2015 | 99,863 (12) |
| 2016 | 102,682 (13) |
| 2017 | 105,905 (13) |
| 2018 | 107,535 (13) |
| 2019 | 108,526 (14) |
| **Year of death among those without hospice enrollment (N=584,414)** |  |
| 2012 | 67,005 (12) |
| 2013 | 67,661 (12) |
| 2014 | 69,614 (12) |
| 2015 | 73,009 (12) |
| 2016 | 74,722 (13) |
| 2017 | 77,250 (13) |
| 2018 | 77,635 (13) |
| 2019 | 77,518 (13) |
| **Year of death among those who discontinued dialysis prior to death without hospice enrollment (N=66,209)** |  |
| 2012 | 7,859 (12) |
| 2013 | 7,637 (11) |
| 2014 | 7,731 (12) |
| 2015 | 8,559 (13) |
| 2016 | 8,454 (13) |
| 2017 | 8,580 (13) |
| 2018 | 8,639 (13) |
| 2019 | 8,750 (13) |
